# “Some alterations in vitamin D metabolism during the COVID-19 pandemic”

**DOI:** 10.1101/2025.07.08.25331143

**Authors:** Dara Dobler, Nicolás A Garrone, Analia I Coralizzi, Eduardo N Cozza Buccaro

**Author notes:** (Corresponding author: Eduardo N Cozza Buccaro,).

## Abstract

Serum levels of vitamin D, calcium and parathyroid hormone during COVID-19 pandemic period (2020-2022) were compared to those of pre-pandemic years (2018-2019) in children, adolescents and older adults. Statistical comparisons showed decreases in vitamin D levels, increases in the proportions of people with serum vitamin D levels below 30 ng/ml (taken as the minimum normal level), decreases in Calcemia, and trend towards increased serum PTH levels in the pandemic period. Within this frame, the COVID-19 pandemic, possibly due to the isolation imposed on the population in Argentina, would have caused alterations in vitamin D metabolism.

## Introduction

### COVID-19 pandemic and outdoor living

The SARS-CoV-2 virus (Severe Acute Respiratory Syndrome Coronavirus 2) is responsible for the disease called COVID-19 (Coronavirus Disease 2019) [1]. The virus and its associated disease have progressively spread around the world, causing the WHO (World Health Organization) to declare a pandemic situation due to COVID-19 in 2020 [2, 3]. In order to prevent the spread of infections (Ro, basic reproduction number) [4], many countries decided to restrict the movement of their inhabitants which was manifested itself in the confinement and quarantine of a large part of the population [2]. In Argentina, this was the goal of the Preventive Social and Mandatory Isolation (APSO) [5].

In this way, a large part of the population reduced outdoor and sun exposure times by increasing the time spent at home [6, 7] during 2020-2022.

### Vitamin D, health and sunlight

Vitamin D is essential for maintaining calcium and phosphorus homeostasis, it plays an important role in the process of bone mineralization through the stimulation of intestinal absorption of calcium from food and the incorporation of calcium into the bones, preventing rickets and osteomalacia [8]. But vitamin D also has multiple effects beyond the regulation of calcemia and bone function, such as its involvement in the regulation of muscle function, cell growth, the immune system, and gene expression, being related to the onset of diabetes mellitus, cancer, autoimmune diseases, metabolic disorders and cardiovascular disease [9, 10]. Thus, vitamin D deficiency is a serious public health problem, since its prevalence can also reach pandemic proportions [11].

Most of the vitamin D in the body comes from cutaneous synthesis by the action of sunlight [12, 13, 14], its ultraviolet component, on 7-dehydrocholesterol. The product of this reaction undergoes a first hepatic hydroxylation yielding 25-hydroxyvitamin D and a second hydroxylation in the renal tubule which yields the active form, 1,25-dihydroxy-vitamin D.

As a consequence, low sun exposure is a risk factor [15, 16, 17] for vitamin D deficiency. In this context, and due to the decrease in exposure to the sun’s rays during childhood, it is important to consider that the risk of vitamin D deficiency is higher in children [18].

### Calcemia, Parathormone and Vitamin D

The definition of plasma calcium levels is fundamental for the bone mineralization process. In conditions of hypocalcemia, the secretion of parathormone (PTH) increases, which produces an increase in bone resorption to increase the plasma calcium level and vitamin D synthesis. On the other hand, vitamin D also increases calcemia by stimulating the absorption of the mineral at the intestinal level [19, 20].

Given the influence that vitamin D has on the capacity for optimal calcium absorption and that parathormone (PTH) is also involved in this function of defining calcium levels, the possible effects of confinement during the COVID-19 pandemic on serum calcium and PTH levels were also studied.

## Methodology

Data on the values of some metabolic markers (see below) were collected from Clinical Analysis laboratories according to the following detail:

### Clinical Analysis Laboratories

forty-three laboratories were surveyed from urban areas of the Departments of Morón, Ituzaingó, Merlo, Moreno, La Matanza, Vicente López, San Martín and San Isidro, of the Province of Buenos Aires, Argentina, all belonging to Greater Buenos Aires, the area adjacent to the city of Buenos Aires.

### Laboratory determinations

The data collected from laboratories correspond to the following determinations:

1. Determination of 25-hydroxy-vitamin D (sum of D2 + D3, calciferol). Method: ELISA, Calbiotech kit

2. Determination of total calcium in blood (free calcium + bound calcium). Method: Spectrophotometric, Liquiform kit, Labtest

3. Determination of serum levels of Parathyroid hormone (PTH). Method: ELISA, Tecan, IBL International GmbH

### Age Ranges

the values of the survey belonged to people of the following three age ranges:

1. Girls/boys from 8 to 12 years old
2. Adolescents from 13 to 17 years old
3. Adults over 65 years old

### Periods of the survey determinations

1. Period I: Pre-COVID-19 pandemic: from July 2018 to June 2019
2. Period II: COVID-19 pandemic: from July 2020 to June 2022)

### Anonymity and Identity Protection

the values of the determinations surveyed were carefully protected by the laboratories for ethical purposes as follows:

i. the values surveyed belong to people who signed the authorization for the use of. In many cases this authorization was obtained after the clinical analyses by contacting the patient. This circumtances provoked a large delay in the realization of the study.
ii. the data were collected anonymously for scientific purposes, without revealing the identity of the patient
iii. the information sent by each laboratory only included, in addition to the values of the analyses, the age of the person and the date of the study, without any other data such as gender, weight, address, etc…
iv. in a similar way, the data was protected so that each of the laboratories was unaware of the information sent by another laboratory.
v. in no case it is possible to identify the people who had the determinations whose values were analyzed in this study.

### Statistics

the statistical contrasts shown in this study were performed by Analysis of Variance (ANOVA), within each age range for the two time periods in which the values of the determinations were collected. For the percentages calculated and reported, the contrasts for the difference in proportions were applied.

The sample sizes, whenever possible, were taken equal to or greater than 30 (n ≥ 30) forming larger samples. When this was not possible due to the number of available determinations, it was then assumed that the variable under study follows a normal distribution for the purposes of applying ANOVA.

The difference between the values of the same age range for the two periods was considered significant with a p ? 0.05.

## Results

1. Serum levels of 25-hydroxy-vitamin D (sum of D2 + D3, calciferol) in Children, Adolescents and Adults over 65 years of age

Table 1 shows the serum levels of vitamin D. In all age ranges surveyed, the serum levels of vitamin D are significantly lower in the period 2020 - 2022 compared to the period 2018 - 2019, which would indicate a decrease in vitamin D levels during the COVID-19 pandemic period.

**Table 1.** Serum vitamin D levels (ng 25-hydroxy-vitamin D/ml) in children aged 8 to 12 years, adolescents aged 13 to 17 years, and in adults aged 65 years and older (mean ± standard deviation)

| Pre- COVID-19 pandemic<br>2018 – 2019 |  |  | COVID-19 pandemic<br>2020 – 2022 |  |  |
| --- | --- | --- | --- | --- | --- |
| Ages and sample sizes (n) |  |  | Ages and sample sizes (n) |  |  |
| 8-12<br>n = 35 | 13-17<br>n = 41 | > de 65<br>n= 33 | 8-12<br>n = 30 | 13-17<br>n = 31 | > de 65<br>n= 32 |
| 41,3 ± 6,4 (a) | 34, 7 ± 5,1 (b) | 32,1 ± 4,8<br>(c) | 28,4 ± 6,7<br>(a) | 21,0 ± 4,5 (b) | 20,6 ± 3,9 (c) |

Significant differences (a) $p < 0,01$ ; (b) $p < 0,02$ ; (c) $p < 0,05$

On the other hand, vitamin D values, by period, were not different for the 3 age ranges considered.

Table 2 shows the percentages of decrease in serum levels of vitamin D, by age range, taking for this calculation the averages reported in Table 1

**Table 2.**
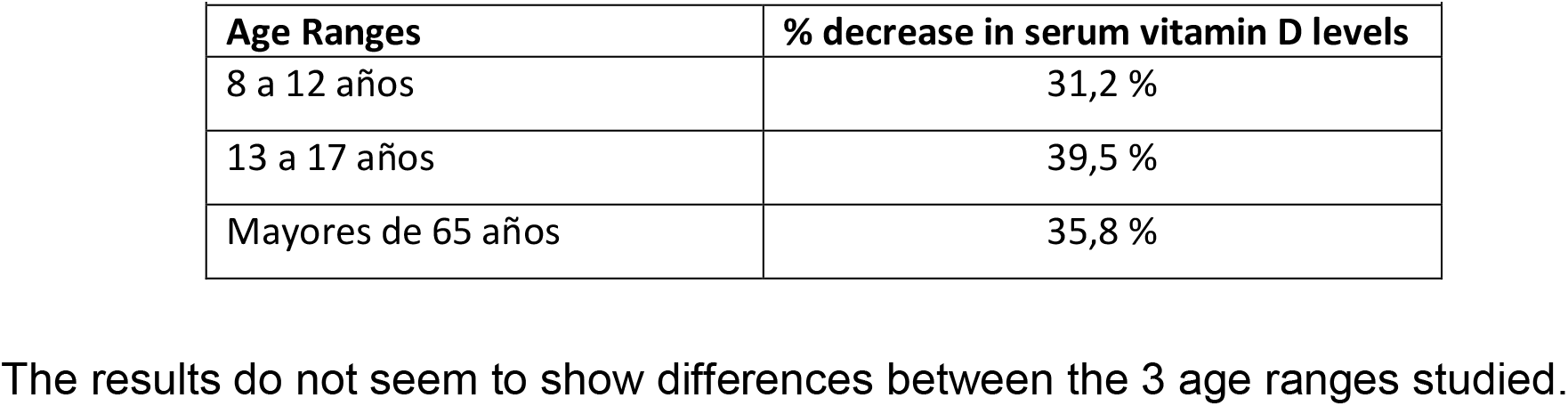
Percentage decrease in serum vitamin D levels in the 2021- 2022 period compared to the 2018-2019 period in the 3 age ranges studied.

| Age Ranges | % decrease in serum vitamin D levels |
| --- | --- |
| 8 a 12 años | 31,2 % |
| 13 a 17 años | 39,5 % |
| Mayores de 65 años | 35,8 % |

2. Deficiency of 25-hydroxy-vitamin D

The accepted minimum normal value for serum vitamin D (sum of D2 + D3, calciferol) is set at 30 ng/ml. Lower values (less than 30 ng/ml) are considered as deficiency of the vitamin. In order to characterize the data collected as sufficient or deficient, the percentages of serum vitamin D values that were lower than the minimum normal value considered here (30 ng/ml) are reported in Table 3.

**Table 3.** Percentages of values with serum vitamin D levels less than 30 ng/ml in children aged 8 to 12 years, adolescents aged 13 to 17 years, and adults aged 65 years and older.

| Pre- COVID-19 pandemic<br>2018 – 2019 |  |  | COVID-19 pandemic<br>2020 – 2022 |  |  |
| --- | --- | --- | --- | --- | --- |
| Ages and sample sizes (n) |  |  | Ages and sample sizes (n) |  |  |
| 8-12<br>n = 35 | 13-17<br>n = 41 | > de 65<br>n= 33 | 8-12<br>n = 30 | 13-17<br>n = 31 | > de 65<br>n= 32 |
| 14,5 % (a) | 36, 7 % (b) | 58,1 % (c) | 48,5 % (a) | 71,2 % (b) | 81,3 % (c) |

Significant differences (a) $p < 0,01$ ; (b) $p < 0,02$ ; (c) $p < 0,05$

These results indicate that for the three age ranges studied, the percentages of people with serum vitamin D levels considered deficient, below 30 ng/ml, increase in the period 2020-2022 compared to the period 2018-2019.

Additionally, the percentages of people with serum vitamin D levels considered deficient tend to increase when moving from the values for Children to those for Adolescents and those for Older Adults (Table 3).

3. Serum levels of total calcium (free calcium + bound calcium)

The results in Table 4 indicate that for the three age ranges surveyed, total blood calcium levels are significantly lower in the period 2020 - 2022 (Pandemic) compared to the period 2018 - 2019 (Pre-pandemic). The percentages of decrease were 13.2, 15.8 and 16.3 respectively for children, adolescents and older adults, respectively.

**Table 4.** Serum levels of total calcium (mg of total Ca2+ / dl) in children aged 8 to 12 years, adolescents aged 13 to 17 years and in adults over 65 years of age (mean ± standard deviation)

3.- Serum levels of total calcium (free calcium + bound calcium)
| **Table 4: Serum levels of total calcium (mg of total Ca<sup>2+</sup> / dl) in children aged 8 to 12 years, adolescents aged 13 to 17 years and in adults over 65 years of age (mean ± standard deviation)** |  |  |  |  |  |
| --- | --- | --- | --- | --- | --- |
| **Pre- COVID-19 pandemic**<br>**2018 – 2019** |  |  | **COVID-19 pandemic**<br>**2020 – 2022** |  |  |
| **Ages and sample sizes (n)** |  |  | **Ages and sample sizes (n)** |  |  |
| 8-12<br>n = 38 | 13-17<br>n = 39 | > de 65<br>n= 37 | 8-12<br>n = 31 | 13-17<br>n = 31 | > de 65<br>n= 30 |
| 9,1 ± 1,3 (a) | 9,5 ± 0,9 (b) | 10,4 ± 1,2(c) | 7,9 ± 1,1 (a) | 8,0 ± 1,2 (b) | 8,7 ± 1,0 (c) |

Significan differences (a): $p < 0.05$ ; (b): $p < 0.05$ ; (c): $p < 0.02$

4. Serum levels of Parathyroid hormone (PTH)

In all the age groups studied, a tendency towards an increase in the values of the variable was evident when moving from the 2018-2019 period to the 2020- 2022 period. The percentages of increase in PTH levels in each age range were 43.7%, 34.5% and 24.9% for children, adolescents and those over 65 years of age respectively. However, these increases were only statistically significant for adolescents (Table 5).

**Table 5.** Serum Parathormone (PTH) levels (pg/ml) in children aged 8 to 12 years, adolescents aged 13 to 17 years and adults over 65 years of age (mean ± standard deviation)

4.- Serum levels of Parathyroid hormone (PTH)
| **Table 5: Serum Parathormone (PTH) levels (pg/ml) in children aged 8 to 12 years, adolescents aged 13 to 17 years and adults over 65 years of age (mean ± standard deviation)** |  |  |  |  |  |
| --- | --- | --- | --- | --- | --- |
| **Pre- COVID-19 pandemic**<br>**2018 – 2019** |  |  | **COVID-19 pandemic**<br>**2020 – 2022** |  |  |
| **Ages and sample sizes (n)** |  |  | **Ages and sample sizes (n)** |  |  |
| 8-12<br>n = 16 | 13-17<br>n = 27 | > de 65<br>n= 21 | 8-12<br>n = 21 | 13-17<br>n = 19 | > de 65<br>n= 16 |
| 46,9 ± 12,3 | 58,2 ± 14,7(a) | 71,0 ± 31,2 | 67,4 ± 20,1 | 78,3 ± 14,2(a) | 88,7 ± 31,4 |

Significan differences (a): $p < 0,05$

## Discussion

Vitamin D deficiency is a global health problem that cannot be underestimated and is associated with a variety of conditions and diseases, not only including bone disorders.

On the other hand, the regulation of blood calcium is subject to a classic endocrine feedback loop [21]. Indeed, calcium from the extracellular fluid binds and activates the calcium-sensing receptor (CaSR) on parathyroid cells, leading to an increase in intracellular calcium that ultimately leads to a reduction in parathyroid hormone (PTH) release [22]. Under hypocalcemic conditions, the calcium fails to bind to its receptor on parathyroid cells leading to the opposite sequence of events, with a consequent increase in PTH production and secretion. This attempts to restore serum calcium levels as PTH increases renal calcium reabsorption and bone resorption by releasing calcium, and also by stimulating renal conversion of 25-hydroxyvitamin D to the active form 1,25- hydroxyvitamin D which in turn increases intestinal calcium absorption and thus calcemia. These effects restore calcium from the extracellular fluid and lead to the inhibition of further PTH production [20]. Within this framework, this data collection was carried out to find out whether the situation of social isolation caused by the COVID-19 pandemic was reflected in any way in the laboratory analyses referring specifically to the endocrine system of vitamin D.

The analysis of the data collected in this study comparing the period 2020-2022 (pandemic) with respect to the period 2018-2019 (pre-pandemic) for the age ranges of children, adolescents and older adults, revealed the following evidence:

i. Decrease in serum vitamin D levels
ii. Increase in the proportion of people with serum vitamin D levels below 30 ng/ml (taken as the minimum normal level)
iii. Decrease in Calcemia
iv. Trend towards an increase in serum PTH levels statistically significant only in adolescents

The metabolic alterations reported in this study could be due to the decrease in sun exposure for long periods of time during the COVID-19 pandemic due to the long and severe confinement and isolation in Argentina. In effect, the low solar radiation on the skin may have led to a decrease in the cutaneous synthesis of vitamin D, which could have led, on the one hand, to a decrease in calcemia and consequently to a tendency towards an increase in PTH, which in our survey was not statistically significant in all three age ranges studied.

The results reported in this study contain the relativity of not having been obtained by paired sample protocols (for example, pre- and post-pandemic data corresponding to the same person), and/or of not coming from a previously established and selected protocol. On the contrary, this survey has begun to be carried out in the final stages of the COVID-19 pandemic, August 2022 in Argentina. The logical ethical provisions of total anonymity, such as signed authorization, provoked a delay in the realization of this study. In this way, data presented here represent statistics of the values reported and only intend to highlight some health issues that would have occurred as an indirect consequence of COVID-19, specifically due to isolation and quarantine.

The existence of hypovitaminosis D together with hypocalcemia has been reported during the COVID-19 pandemic, in severe patients with COVID-19 [23]. On the other hand, given the multiple functions of vitamin D in the body, vitamin D deficiency is a special alert for health authorities.

Based on these results, before similar future health challenges, it is important to design programs and public policies aimed at maintaining and promoting the practice of physical activity, ensuring health checks and encouraging the promotion of healthy eating in all health centers starting from the first level of care, without ruling out the need for vitamin D supplementation [24, 25].

## Data Availability

All data produced in the present study are available upon reasonable request to the authors
All data produced in the present work are contained in the manuscript

## Acknowledgements

The authors would like to thank to Departamento de Ingeniería Química, Facultad Regional Buenos Aires, Universidad Tecnológica Nacional, for creating the conditions to carry out this study and to ILCET for funding support.

